# Modelling the expected and potentially avertable burden of infection-related cancer among newly arrived migrants in the European Union

**DOI:** 10.64898/2026.10.28.26366414

**Authors:** Rachel Wittenauer, Damien Georges, Jin Young Park, Andrea Gini, Irene Man, Freddie Bray, Stefano Rosso, Iacopo Baussano, Catharina Johanna Alberts, the Cancer RADAR working group

**Affiliations:** International Agency for Research on Cancer (IARC/WHO), Early Detection Prevention and Infections Branch, Lyon, France; International Agency for Research on Cancer (IARC/WHO), Cancer Surveillance Branch, Lyon, France; Piedmont Cancer Registry, University Hospital Città della Salute e della Scienza di Torino and CPO Piemonte, Turin, Italy; Epidemiology and Data Science Department, Amsterdam UMC, and Public Health Service of Amsterdam, Amsterdam, the Netherlands

## Abstract

International migration is evolving globally. Newly arrived migrants (individuals relocating into countries in the European Union) are a European Centre for Disease Prevention and Control (ECDC) priority population for entry health assessments, offering opportunities to prevent and treat cancer-associated infections, although implementation varies across Europe. Using migration, cancer incidence, and intervention data, we modelled the burden of hepatocellular carcinoma (HCC), cervical cancer (CvC), and gastric cancer (GC) and the proportion avertable by HBV vaccination, HBV and HCV screen-and-treat, HPV vaccination, cervical screening, and *Helicobacter pylori* screen-and-treat. We estimated 636,000 lifetime cancer cases (145,000 HCC, 155,000 CvC, 336,000 GC). Assuming full uptake, among eligible migrants, ECDC-recommended HBV vaccination, HBV screen-and-treat, and HCV screen-and-treat could avert 36%, 26%, and 17% of expected HCC cases, respectively; HPV vaccination and cervical screening 65% and 60% of CvC cases, respectively; and *H. pylori* screen-and-treat 57% of GC cases. Expanding eligibility could further increase the impact.

## Introduction

Migration patterns are diversifying globally, shaped by economic inequalities, climate change, conflict, and other complex dynamics (1). In 2024, nearly 5 million new international migrants entered countries in the European Union (EU), corresponding to 1.1% of the population (2–4). While some aspects of migration patterns vary across regions, broad global trends include ageing populations, complex health burdens among migrants, and constrained healthcare systems. Support for migrants upon entry is essential, as those newly arrived (defined here as individuals arriving in any country in the EU in the past five years, including both intra-EU and extra-EU migrants) frequently encounter structural barriers to care and interruptions in preventive health services (3–5). Initial health assessments upon arrival offer an important opportunity to identify health needs, re-establish continuity of care, and provide preventive services missed before or during migration, consistent with European Centre for Disease Prevention and Control (ECDC) recommendations (3, 6).

Current global and regional policy guidance for preventive health services for newly arrived migrants, such as from the World Health Organization (WHO) and the ECDC, primarily focuses on communicable diseases of immediate transmission potential, such as tuberculosis, HIV, hepatitis B and C, and vaccine-preventable infections (4–6). However, these recommendations rarely address the longer-term impact of these infections on noncommunicable diseases (NCDs), including cancer, and generally exclude the two infectious diseases that worldwide contribute to the largest burden of infection-related cancers, namely *Helicobacter pylori* (*H. pylori*) and human papillomavirus (HPV) (7). Integrating prevention of these long-term risks into migrant health guidance could reduce the future cancer burden within the EU (8–10).

Information on the scale and profile of cancer among migrants remains fragmented and primarily composed of smaller-scale studies in specific sub-populations (5, 11, 12). Current evidence indicates that migrants have an elevated risk of infection-related cancers relative to their host populations (11, 13). Worldwide, the four pathogens responsible for 88% of the infection-related cancer burden are hepatitis B (HBV), hepatitis C (HCV) (both causing hepatocellular carcinoma i.e. liver cancer), human papillomavirus (HPV) (causing several cancers, of which cervical cancer is the most important), and *H. pylori* (causing several cancers, of which gastric cancer is the most important) (7, 14). Infectious causes of cancer are highly modifiable risk factors which lend themselves well to treatment and prevention services, namely HBV vaccination; HBV and HCV test-and-treat programs, *H. pylori* screen-and-treat programs; HPV vaccination; and cervical cancer screening (14, 15).

Although migration is a global phenomenon, the EU provides a unique context for this analysis because of the (i) existing region-wide guidance for migrant health assessments, (ii) availability of up-to-date data on newly arrived migrants, and (iii) wide geographic variation in cancer burden and resources for health among EU member states. Despite the availability of high-quality guidance from the ECDC (6) and robust demographic data from Eurostat (16), the infection-related cancer burden among newly arrived migrants remains understudied. This is partly due to limited data on migrants’ post-migration cancer risk and substantial uncertainty in care cascade parameters across countries of origin and destination. However, without a comprehensive understanding of the infection-related cancer burden among migrants and potential impact of preventive interventions, migrant health policies are likely to remain insufficient, as they cannot be appropriately targeted, prioritised, or scaled.

The objectives of this study are to: (1) estimate the expected number of hepatocellular carcinoma (HCC), cervical cancer (CvC), and gastric cancer (GC) cases among migrants arriving in EU countries over the most recent five years for which data are available (hereafter referred to as “**expected cases**”); (2) quantify the infection-attributable proportion of these cancers (hereafter referred to as “**infection-attributable cases**”); and (3) estimate the potential impact of implementing preventive interventions at entry (hereafter referred to as “**potentially averted cases**”). **Potentially averted cases** were examined in three steps: (3a) implementation of **existing EU guidance** for newly arrived migrants which currently focuses on hepatitis prevention and treatment; (3b) expansion of guidance to include HPV vaccination, cervical screening, and *H. pylori* screen-and-treat (“**expanded guidance**”); and (3c) broadening eligibility criteria for the hepatitis interventions and *H. pylori* screen-and-treat (“**eligibility scenarios**”).

## Results

There were approximately 22 million persons who migrated into EU countries over the most recent five years with available data (2). Hereafter, ‘migrants’ refers to people who migrated into countries of the EU27 (including both intra-EU and extra-EU migrants) over the period 2020-2024. On average, the migrants were 54% men and 46% women with a median age of 29 years (IQR: 20-40). The ongoing displacement from Ukraine is reflected in our data: across the 27 host countries, the median proportion of migrants from Ukraine was 11.1% (IQR: 4.4 – 19.5%), with the highest proportion in Czechia (75% from Ukraine), and with a total number of Ukrainian migrants peaking in 2022. The impact of the COVID-19 pandemic travel restrictions is also reflected in our data: in 2020, the total number of migrants was 2.6 million, and in 2021 the total migrants returned to 2019 pre-pandemic levels of approximately 3.3 million (***Supplementary Figure S-1***). The host countries with the highest number of total migrants were Germany (5.5 million), Spain (4.5 million), France (1.7 million), and Italy (1.5 million). By continent of birth, migrants were most often from within Europe (50%, primarily Eastern Europe (32%)), followed by Asia (18%), Africa (14%), Latin America (14%), Northern America (1.5%), Oceania (<1%), and unknown origin (2%). Regarding cancer incidence in the country of birth, the proportion of migrants born in a country with an ASR above the global median of country-level ASRs was 29% for HCC, 46% for CvC and 65% for GC. (***Figure 1***).

**Figure 1.**
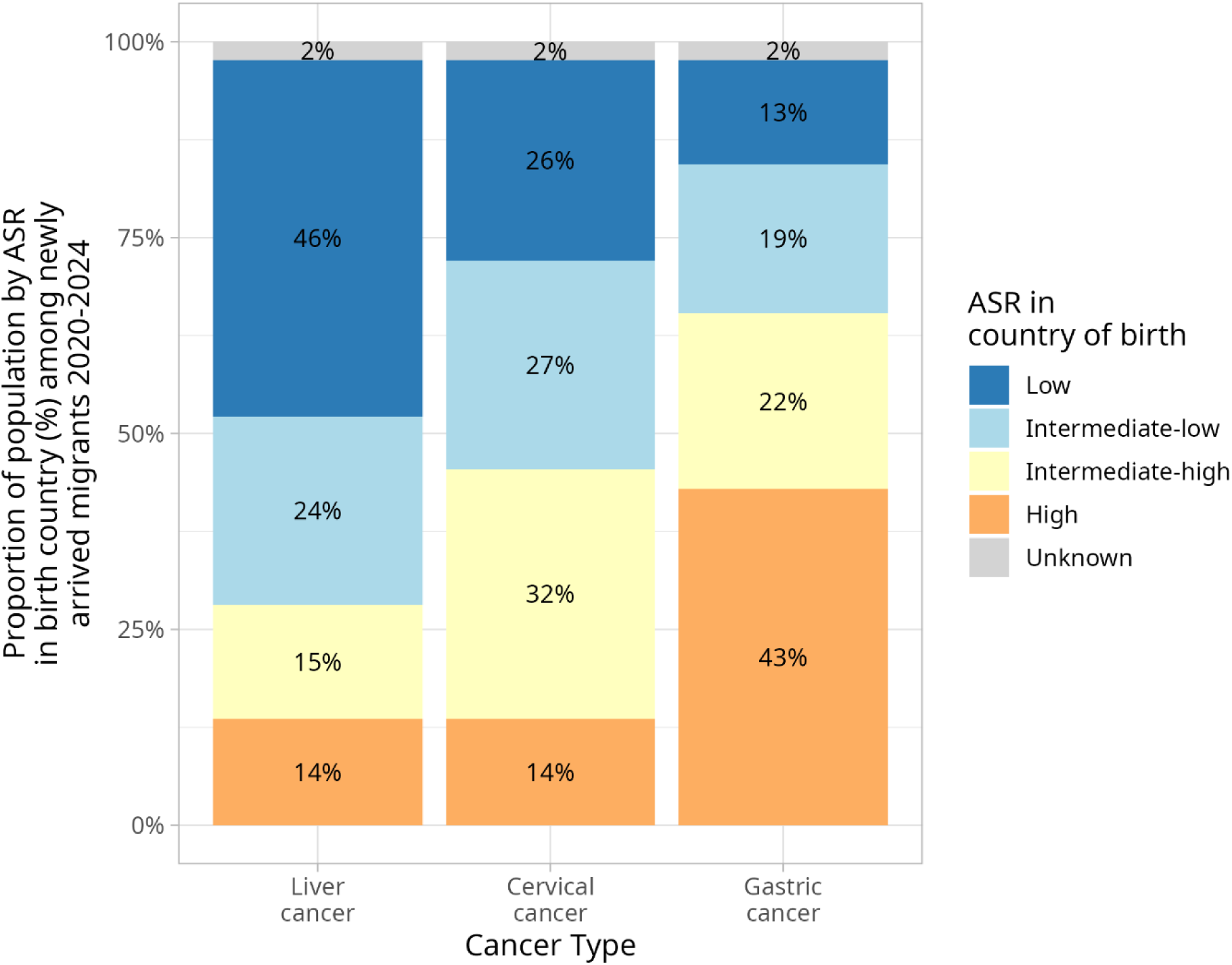
Proportion of migrants arriving in the EU27 in 2020–2024, by quartile of the worldwide age-standardized incidence rate (ASR) of liver cancer, cervical cancer and gastric cancer in their country of birth. Colours refer to the ASR in the birth country of the migrant. The grouping was based on the quartile of the worldwide distribution of ASR of the respective cancer under consideration. ASR categories (per 100 000 person-years, both sexes) corresponded to the quartiles of the worldwide distribution: **liver (quartiles available for liver cancer, not specifically HCC)**, low <2.51, intermediate-low 2.51-<3.75, intermediate-high 3.75-<6.01, high ≥6.01; cervical cancer, low <8.02, intermediate-low 8.03-<13.81, intermediate-high 13.81-<21.51, high ≥21.51; **gastric cancer**, low <3.73, intermediate-low 3.74-<5.51, intermediate-high 5.51-<9.39, high ≥9.39. Unknown indicates migrants with missing country of birth. May not sum to 100% due to rounding. **Source data:** Globocan 2024 and Eurostat 2020-2024 (10, 16).

### Expected total and infection-attributable cancer cases

Regarding *expected cases* (objective 1), among 5 years of newly arrived migrants, we estimated that 635,542 infection-related cancer cases will arise over their lifetime in the absence of additional preventive interventions. This total comprised 144,618 HCC (23%) cases, 154,688 CvC cases (24%), and 336,235 GC (53%) cases. By region of birth, 302,259 (48%) of these cases were among people born in Europe, of whom 142,239 (22% of the total cases) were born in EU27 countries (***Figure 2***). Of the 635,542 total cases, 93,583 (15%) were among migrants from Ukraine.

**Figure 2.**
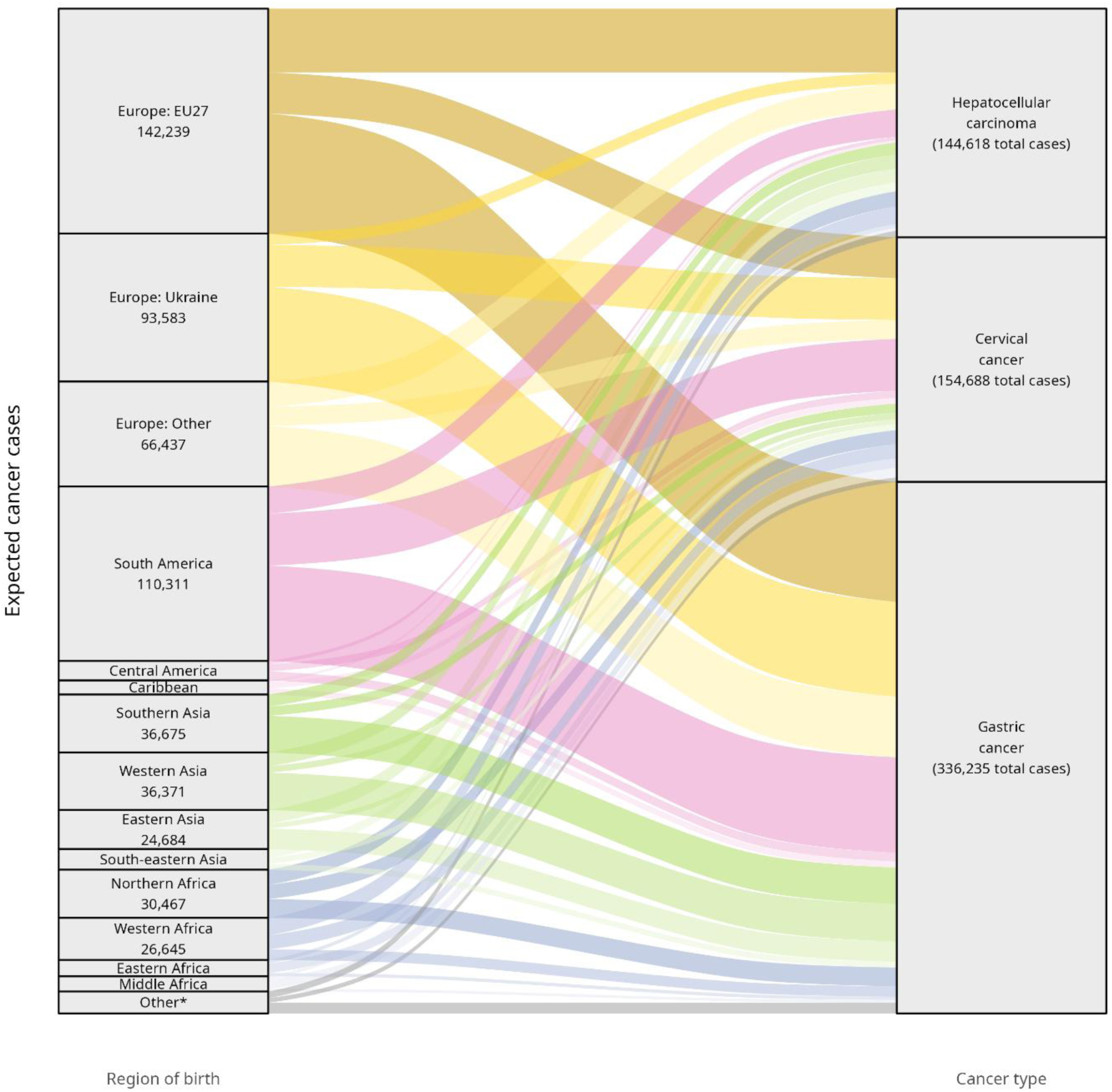
Expected cancer cases among migrants arriving in the EU27 in 2020–2024, by UN subregion of birth and cancer type (hepatocellular carcinoma, cervical cancer, gastric cancer). Within the European region, the darkest yellow denotes EU27 countries, the medium yellow denotes Ukraine, and the lightest yellow denotes all other countries in Europe. The “Other*” countries comprise the birth countries which make up less than 1% of expected cancer cases: Northern America (0.9%), Central Asia (0.8%), Southern Africa (0.3%), Australia and New Zealand (0.2%), Melanesia (<0.1%), and Polynesia (<0.1%).

By sex, 309,350 (49%) of these cases were among women (40,093 HCC, 154,688 CvC, and 114,568 GC cases), and 326,192 (51%) among men (104,525 HCC cases and 221,667 GC cases) (***Figure 3.A***). Among HCC cases, 57% were among migrants born in countries with an ASR above the global median and for CvC and GC this proportion was 65% and 85%, respectively (***Figure 3.B***). The host countries with the highest estimated total cancer burden among newly arrived migrants were Germany and Spain (see ***Supplementary Figure S-2***).

**Figure 3.**
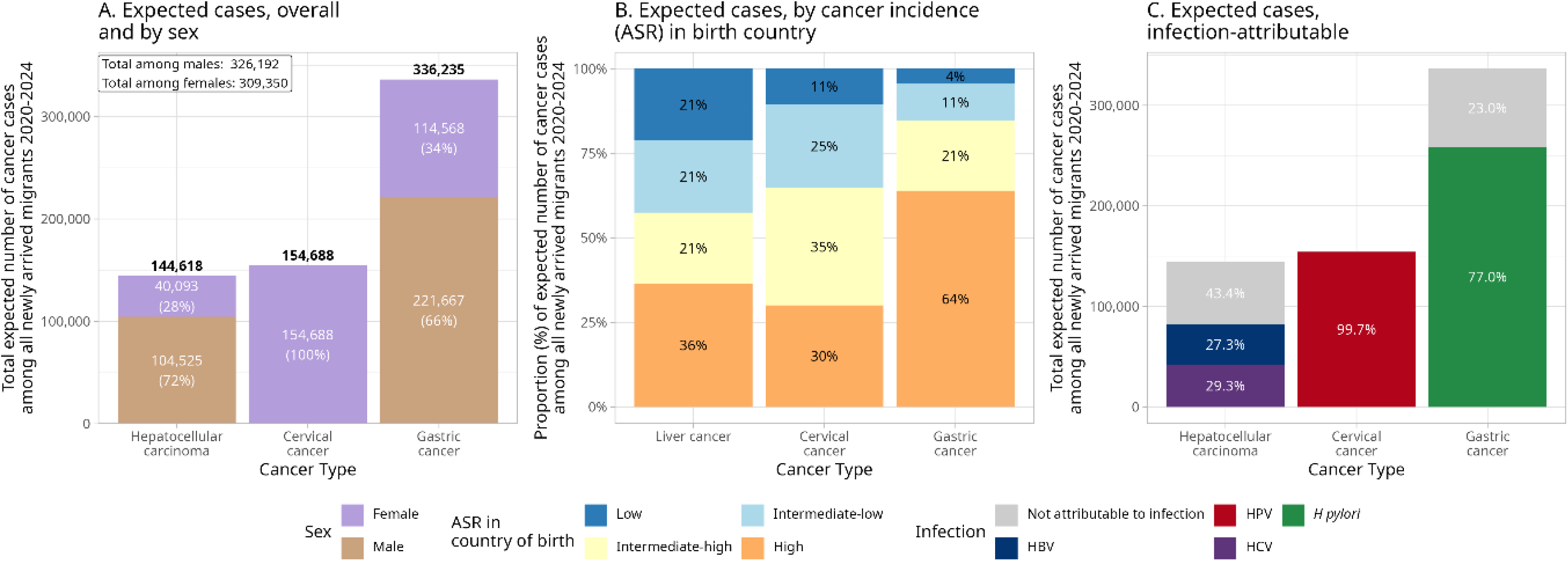
The expected lifetime number of hepatocellular carcinoma, cervical cancer, and gastric cancer cases among migrants arriving in EU27 countries in 2020-2024, by sex, incidence levels in the country of birth, and attribution to infection. Expected cases are cumulative over the remaining lifetime of migrants (up to 80 years after arrival). **Panel A** disaggregates the totals by sex of migrants. **Panel B** stratifies the totals by quartiles of the worldwide distribution of age-standardized incidence rates (ASR) for the respective cancer in the birth country. Low ASR (dark blue) corresponds to migrants from low-incidence countries, and high ASR (orange) corresponds to migrants from high-incidence countries of the respective cancer. See Figure 1 for the ASR values corresponding to each quartile. **Panel C** highlights the proportion of each cancer type attributable to the corresponding infection (“infection-attributable cancer cases”, namely HBV or HCV for hepatocellular carcinoma, HPV for cervical cancer, and H. pylori for gastric cancer), which is the proportion targeted by the interventions modelled in this analysis. **Abbreviations:** ASR: age-standardized incidence rate; HBV: hepatitis B virus; HCV: hepatitis C virus; HPV: human papillomavirus.

Regarding *infection-attributable cases* (objective 2), of the total, 494,946 (78%) cases were attributable to the infections under consideration: 81,875 HCC from HBV/HCV (57%), 154,224 CvC from HPV (99.7%), and 258,847 GC from *H. pylori* (77%) (***Figure 3.C***).

### Strategies for prevention

Potentially averted cases (objective 3) are presented for the three steps: (3a) *current EU guidance* on hepatitis, (3b) *expanded guidance* including HPV and *H. pylori* interventions, and (3c) broader *eligibility scenarios*.

#### 3.a Current EU guidance: strategies for HCC prevention

We estimated that full implementation of the current ECDC viral hepatitis recommendations which target migrants born in countries with HBV or HCV prevalence ≥2% (Table 1), would reduce the HCC burden by 15% (21,520 averted in total by the three interventions, out of 144,618 cases) among all migrants. By individual intervention, HBV vaccination, HBV screening and treatment, and HCV screening and treatment were estimated to avert 36% (3,425 cases), 26% (14,420), and 17% (3,675), respectively, of all expected HCC cases among migrants eligible for each intervention under ECDC criteria (***Figure 4***). The remaining cases were either attributable to HBV or HCV but not averted, or not attributable to viral hepatitis (grey in Figure 4) and therefore cannot be averted by these interventions.

**Figure 4.**
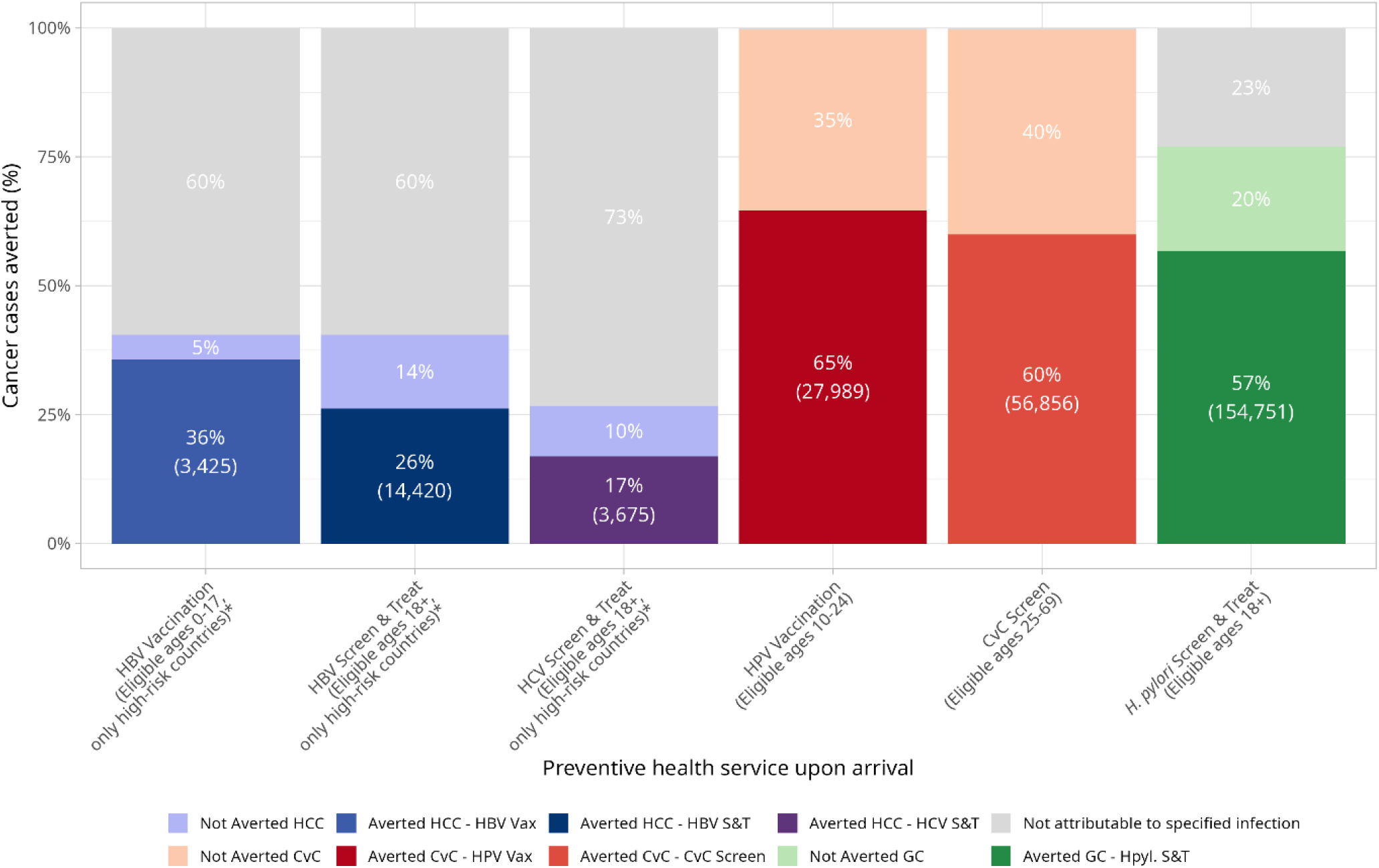
Proportion of expected lifetime cancer cases averted by each intervention among eligible migrants arriving in the EU27 in 2020-2024. Expected cases are cumulative over the remaining lifetime (up to 80 years after arrival). Each bar represents all expected cancer cases of the respective cancer among migrants eligible for the intervention (100%), divided into cases averted by the intervention (darker shade), infection-attributable cases not averted (lighter shade), and cases not attributable to infection (grey), which cannot be averted by interventions evaluated. Eligible populations differ between interventions, so bars are not additive. Averted cases reflect the modelled effectiveness of each intervention (see Table 1). *High-risk countries are defined according to ECDC recommendations as countries with an HBV prevalence of ≥2% (for HBV vaccination, ages 0–17 years, and HBV screen-and-treat, ages ≥18 years) or an HCV prevalence of ≥2% (for HCV screen-and-treat, ages ≥18 years) (39). **Abbreviations**: CvC: cervical cancer; GC: Gastric cancer; HBV: hepatitis B virus; HCC: hepatocellular carcinoma; HCV: hepatitis C virus; HPV: human papillomavirus; S&T: screen-and-treat.

**Table 1.** Modelling approach and key parameters by cancer type and intervention.

| **Cancer** | **Liver (hepatocellular carcinoma (HCC))** |  |  | **Cervical** |  | **Gastric** |
|---|---|---|---|---|---|---|
| **Preventive health service** | Vaccination for HBV | Screening for HBV and treatment (ETV or TDF) | Screening for HCV, RNA confirmatory, treatment (DAAs) | Vaccination for HPV (nonavalent vaccine) | Testing for HPV, corresponding CvC screening follow up | Screening for *H. pylori* with urea breath test and 14-day bismuth quadruple therapy for treatment |
| **Currently recommended strategy to be actively offered for newly arrived migrants (6)** | Yes (obj.3a) | Yes (obj.3a) | Yes (obj.3a) | No (obj.3b) | No (obj.3b) | No (obj.3b) |
| **Eligibility** | Men & women ages 0-17 from countries with HBV prevalence ≥ 2% (39);<br>Eligibility varied in obj.3c | Men & women ages 18+ from countries with HBV prevalence ≥ 2% (39) ;<br>Eligibility varied in obj.3c | Men & women ages 18+ from countries with HCV prevalence ≥ 2% (39);<br>Eligibility varied in obj.3c | Girls and women ages 10-24 from all countries (40) | Girls and women ages 25-69 from all countries (41) | Men and women ages 18+ from all countries (42) ;<br>Eligibility varied in obj.3c |
| **Modelled Effectiveness** | 95% reduction in HBV-attributable HCC incidence ages 0-14, 85% for 15-17 (43) | 65% effective preventing HCC (44-46) | 65% effective preventing HCC (47, 48) | 95% effective against HR-HPV types (total AF for 9-valent 95.7% (49)) for ages 10-14, 85% for 15-19, and 50% for 20-24 (50, 51) | 60% effective preventing cervical cancer (52) | 80% effective in 18-39; 60% effective in ages 40+ (42, 53) |
| **Modelled attributable fraction of agent on cancer** | Country-specific, median 34% of HCC attributable to HBV (14, 54) | Country-specific, median 34% of HCC attributable to HBV (14, 54) | Country-specific, median 24% of HCC attributable to HCV (14, 54) | 99.7% of CvC attributable to HPV (55) | 99.7% of CvC attributable to HPV (55) | 89% non-cardia GC and 0% cardia GC (except in Eastern Asia excluding China (29%) and China (62%)) attributable to *H. pylori* (14, 42) |
| **Modelled Incidence** | Age- and sex-specific IR of birth country (10) | Age- and sex-specific IR of birth country (10) | Age- and sex-specific IR of birth country (10) | Age- and sex-specific IR of birth country (10) | Age- and sex-specific IR of birth country (10) | Age- and sex-specific IR of birth country (10) |

**Abbreviations:** AF: Attributable fraction; CvC: Cervical cancer; DAA: Direct acting antivirals; ETV: entecavir; IR: Incidence rate; GC: Gastric cancer; HBV: Hepatitis B virus; HCC: Hepatocellular carcinoma; HCV: Hepatitis C virus; HPV: Human papillomavirus; HR: High-risk; obj.3a: objective 3 a; obj.3b: objective 3b; obj.3c: objective 3c; TDF: tenofovir

#### 3.b Expanded guidance: strategies for CvC and GC prevention

Among migrants eligible for the interventions, HPV vaccination (women aged 10-24 years) and cervical screening (women aged 25-69 years) were estimated to avert 65% and 60% of expected CvC cases, respectively (***Figure 4***). ***Supplementary Figure S-3*** provides a further breakdown of expected and potentially averted cases by age group among all migrants.

*H. pylori* screen-and-treat offered to adult migrants would potentially avert an estimated 46% (154,751 out of 336,235 cases) of GC cases among all migrants. Among migrants eligible for the intervention, an estimated 57% of GC cases would be averted (***Figure 4***), while 20% of GC cases are attributable to *H. pylori* but would not be averted (light green), mainly because screen-and-treat is not assumed to be 100% effective (***Table 1***). Approximately 23% of cases among eligible migrants are not attributable to *H. pylori* (grey) and therefore cannot be averted by this intervention.

#### 3.c Eligibility scenarios

For HCC, extending eligibility for the hepatitis interventions to migrants born in countries in the top 25% or top 50% of liver ASR, in addition to the current ECDC prevalence-based criteria (***Table 1***), increased the number of potentially averted HCC cases from 21,520 under current ECDC criteria by 41% (to 30,390) and 79% (to 38,420), respectively. If all migrants were eligible regardless of birth country, potentially averted HCC cases would increase by 132% (to 49,851) (***Figure 5.A***).

**Figure 5.**
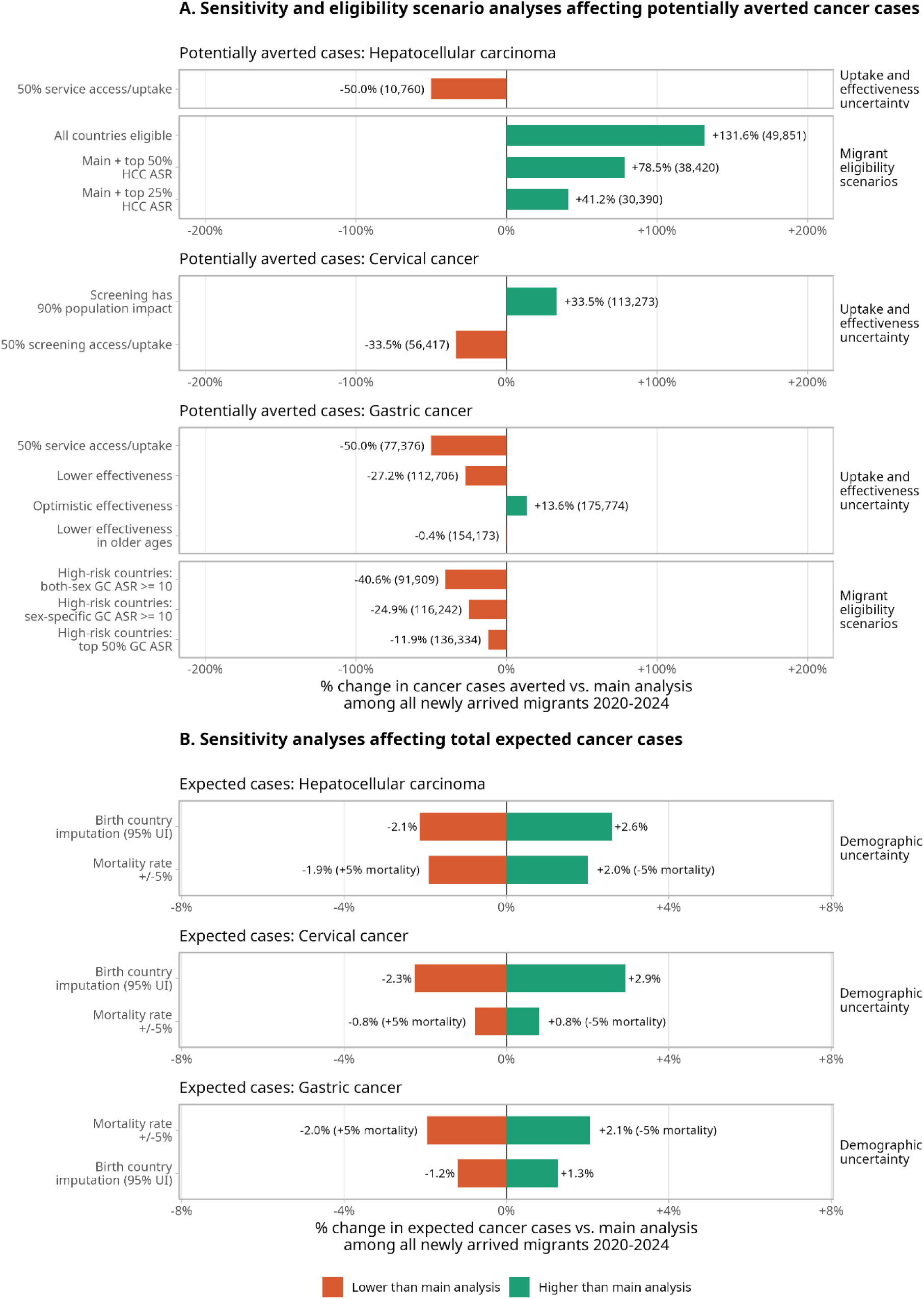
One-way sensitivity analyses and eligibility scenarios of the expected and potentially averted burden. Each key assumption is varied across a plausible range while the others are held at the reference value. panel A shows the impact of cancer-prevention-specific parameters (intervention effectiveness, low service uptake, and varying intervention eligibility criteria). Panel B shows the impact of uncertainty for demographic parameters (mortality rate and imputed birth country) which affect the expected number of cancer cases. Bars show change relative to the reference analysis. **Abbreviations:** ASR: age-standardized incidence rate; HCC: hepatocellular carcinoma; GC: gastric cancer; UI: uncertainty interval.

For GC, restricting *H. pylori* screen-and-treat to migrants born in higher-incidence countries reduced the number of potentially averted GC cases compared with offering it to all adult migrants using three different eligibility criteria (***Figure 5.A***). Restricting eligibility to countries in the top 50% of GC ASRs reduced potentially averted cases by only 12% (136,334 averted, versus 154,751 in the main analysis), whereas applying a GC ASR threshold of ≥10 per 100,000 person-years reduced potentially averted cases by 25% (116,242 averted) when based on sex-specific ASRs and by 41% when based on both-sex ASRs (91,909 averted).

#### Sensitivity analyses

We conducted a priori sensitivity analyses to assess how uncertainty in demographic parameters (mortality rates and birth country) and in intervention effectiveness and uptake affected the results (**Figure 5**).

Potentially averted cases were most sensitive to access and uptake (***Figure 5.A***). Assuming 50% access/uptake reduced potentially averted cases by 50% for HCC (to 10,760) and GC (to 77,376). For CvC, assuming 50% access/uptake of screening reduced the total potentially averted CvC cases by 34% (to 56,417).

Assumptions about intervention effectiveness generally had a smaller effect. Increasing the population-level impact of cervical screening from 60% to 90% increased the total potentially averted CvC cases by 34% (to 113,273) (***Figure 5.A***). For *H. pylori* screen-and-treat, assuming lower or optimistic effectiveness changed potentially averted GC cases by −27% and +14%, respectively, whereas lower effectiveness in older ages had a negligible effect (−0.4%) (***Figure 5.A***).

Uncertainty in demographic parameters had minimal effect on the expected cases, changing them by less than 3% for all three cancers (range −2.3% to +2.9%; **Figure 5.B**). Further sensitivity analysis results are provided in ***Supplementary Note A.2***.

## Discussion

Among the approximately 22 million migrants who arrived in EU countries over the most recent five years for which data were available (2020–2024), we expect approximately 636,000 total cases of hepatocellular carcinoma (HCC), cervical cancer (CvC), and gastric cancer (GC) to arise over the next 80 years in the absence of any additional prevention interventions: 145,000 HCC cases, 155,000 CvC cases, and 336,000 GC cases. We evaluated the population-level impact of ensuring full uptake of six infection prevention and treatment services among eligible migrants upon arrival in their host country to prevent cancer.

### Interpretation of results

Each of the six interventions is highly effective in reducing cancer risk at the individual level, so our results as presented in this analysis (impact on cancer burden across the total newly arrived migrant population) might be perceived as low. However, our estimates reflect a combination of (i) migrants who are eligible for the service (which varies by country of birth, age, and sex), (ii) attributable fractions of each infectious agent on the cancer type, and (iii) the intervention effectiveness (which for some decreases with age). For example, although HBV vaccination is highly effective (≈90%), only a proportion of HCC cases are attributable to HBV (global median 34% [IQR 13–54%]), which limits the achievable population-level impact. In addition, in this study, we estimated prevention services upon entry in the host country. As migrants integrate into the host country’s health system, the appropriate prevention or treatment services should be provided in accordance with current healthcare guidelines and age-based eligibility criteria. For example, migrants below the age of HPV vaccination eligibility at arrival were not included in our estimates of averted cases, but are likely to be offered HPV vaccination later through host-country programs. Ensuring equitable access to routine host-country healthcare for migrants could therefore avert additional cases beyond those estimated here. Further, migrants from other EU countries may have already received relevant preventative services in their country of birth or may access them in another EU country after migration, though coverage varies widely throughout the EU and birth countries (17). As our analysis does not account for individual healthcare histories or subsequent migration, we cannot determine where these services are received. Therefore, the estimated cancer burden and potentially avertable cases are reported at the aggregate EU-level, without attributing service delivery or associated benefits to a specific EU country.

### Implications for policy

These results support the potential addition of HPV vaccination, cervical screening, and *H. pylori* screen-and-treat to the current ECDC guidance for newly arrived migrants (6). The number of expected cases and potentially averted cases of CvC (among women) and GC (among men and women) are greater than those of HCC, and could be effectively prevented with services of similar complexity to the viral hepatitis-targeting interventions. The current guidance for viral hepatitis would avert approximately 15% of HCC cases, while HPV vaccination, cervical screening, and *H. pylori* screen-and-treat would avert about 49% of total expected GC and CvC cases (239,596 prevented combined out of 490,923 expected). Notably, GC accounts for the largest share of the expected cancer burden (53%), particularly among men (68% of total expected cancer cases among men and 37% among women). The national adoption of population-wide *H. pylori* screen-and-treat programs may be low-priority in EU countries with low *H. pylori* endemicity. However, screening newly arrived migrants for *H. pylori* offers a unique opportunity to avert a substantial number of cancer cases. There is currently no established definition of which migrants should be considered “high-risk” or eligible for such intervention (18–20). We therefore evaluated universal eligibility (all migrants aged ≥18 years) alongside several risk-based eligibility scenarios based on birth country (similar to viral hepatitis screening criteria). Restricting *H. pylori* screen-and-treat to only migrants from birth countries with GC ASR above the global median resulted in only 12% fewer potentially averted GC cases compared with universal eligibility, whereas a fixed threshold of GC ASR ≥10 per 100 000 person-years resulted in 25% (sex-specific ASR) to 41% (both-sex ASR) fewer averted cases. This suggests that a risk-stratified approach targeting migrants born in countries with GC ASR above the global median (i.e., the top 50% of countries worldwide) could achieve most of the benefit of universal screening while targeting a smaller migrant population.

In addition to these new interventions, policymakers should also consider the expansion of current eligibility criteria for hepatitis-targeting interventions beyond the 2% viral prevalence threshold (6). We propose expanding eligibility criteria to include additional countries based on HCC ASRs, because as countries approach viral hepatitis elimination, current viral prevalence may underestimate historical exposure and future HCC risk. If migrants from high-ASR countries (defined as higher than the global median) were also eligible, about 1.8 times as many HCC cases would be averted (21,500 (15%) vs 38,400 (27%)). Although distribution-based thresholds shift with changing global epidemiology, this is also true for current ECDC eligibility criteria. As general-population prevalence declines and the world moves towards HBV, and eventually HCV, elimination, a prevalence-based threshold would increasingly exclude migrants from targeted intervention, despite a substantial number of individuals with existing infection remaining at risk for progression to liver cancer. Because HCC ASRs reflect historical rather than current infection, eligibility criteria based on birth cohort-specific IRs may better capture these migrants. Prevalence declines first in younger cohorts protected by infant HBV vaccination, whereas migrants born before national HBV vaccination was introduced remain at higher risk. Although this was beyond the scope of our analysis, it warrants further evaluation.

Finally, the capacity of each host country’s healthcare system to provide additional services must be assessed and sufficient resources allocated to implement these services effectively, particularly for hepatitis prevention and *H. pylori* screen-and-treat services (as HPV prevention services are already widely recommended for the general population). Beyond deciding whether to provide these services to newly arrived migrants, the equity impacts and trade-offs between narrowing (fewer cancer cases averted, but fewer resources needed) or expanding eligibility criteria (more cancer cases averted, but more resources needed) for HCC and GC-preventing services will need to be carefully assessed before implementation. Our analysis presented here provides a starting point to inform these policy decisions and guide further implementation considerations.

### Implementation considerations

Migrants often remain underserved after arrival due to multiple structural and practical barriers to accessing healthcare (3–6, 21, 22), and implementation of migrant-specific health policies also face many operational and financial challenges (5). As seen in our sensitivity analyses, access to these services was one of the most influential parameters on number of cancer cases averted. However, evaluations of existing infectious disease screening programs for migrants have been shown to be promising, reporting high acceptability among both patients and service providers (5, 23–25). In this analysis, we optimistically assume that all migrants will access services and complete the appropriate follow-up, but even in sensitivity analyses where we conservatively assume that only 50% of migrants will do so, we found a large proportion of cancer cases could be averted. Offering basic preventive services upon arrival could improve uptake among migrants not yet linked to regular primary care providers (26–28), as is currently the norm with some infectious diseases such as tuberculosis, though programs vary widely across the EU (29).

After migrants arrive and are integrated into routine health systems, providing targeted services will remain essential for comprehensive and sustainable cancer prevention (3, 30). A study of nearly 900 undocumented migrants and refugees in Italy found no HBV vaccination uptake despite a median stay of nearly 5 years (31). Prevention must also address non-infectious risk factors for liver cancer (e.g., diet, alcohol, or metabolic risk factors) as only 57% of HCC cases were attributable to HBV or HCV. Implementation of *H. pylori* screen-and-treat programs presents additional challenges, as these are not routinely implemented in EU countries. Eligibility criteria and measures to mitigate the risk of antimicrobial resistance from widespread antibiotic use, including resistance surveillance, stewardship plans, and protocols for confirmation-of-cure testing, must be established before implementation (18, 32). The ongoing European Commission Initiative on Gastric Cancer is addressing these aspects through the development of evidence-based guidelines and quality assurance scheme on *H. pylori* screen-and-treat programs for Europe (33).

### Strengths and limitations

Our analysis has several strengths. To our knowledge, this is the first study to provide a cancer perspective on prevention of infections among newly arrived migrants by quantifying the infection-related cancer burden. We have used the most up-to-date and detailed data available on migrant population size, birth and host country distributions, and age distributions, alongside cancer incidence by type, sex, age group, and country. We estimate the cancer burden both with and without key prevention services using a publicly available model (34, 35) and data sources, combined with a novel analytic approach that can be replicated for other cancers and settings.

Our analysis also comes with important limitations. The data and assumptions used throughout our analysis, while well-grounded, should be interpreted with caution. Health data stratified by migration background are limited, necessitating several assumptions about risk. Although we focus on newly arrived migrants and use birth-country ASR as a reasonable proxy, detailed data on HBV, HCV, *H. pylori*, or HPV infection are unavailable, limiting the use of more detailed microsimulation models and thus risk-stratified or intervention-specific assumptions. As a result, while based on published evidence, our estimates of the population-level impact of each intervention may not fully capture the complexities of each intervention. As individual histories of preventive interventions before migration are unavailable, we estimate the potential preventable cancer burden among migrants who arrived in EU countries within the last five years. More detailed individual-level data could further refine these estimates. Our analysis focuses on population-level prevention potential rather than health-economic evaluation, cost-effectiveness, cost-utility, or budget-impact analysis. Treatment costs, health-system capacity to absorb cases, and budget impact are nonetheless important for implementation and are best addressed through dedicated economic evaluations, which fall outside the scope of the study.

We applied host-country all-cause mortality to capture the competing risk of mortality. However, migrants’ mortality may differ from, and in some groups exceed, host-population mortality, which would reduce time at risk and therefore overestimate the expected number of cancer cases. We do not model onward or return migration, as reliable data on how often a newly arrived cohort leaves, or where those who leave go, are lacking. Many migrants who move may remain within the EU, where they would continue to benefit from prevention offered at entry.

Birth-country and age detail were imputed for a minority of host countries, but sensitivity analyses suggested that the impact of these imputations on the estimates was limited. We base our analysis on migrants who arrived within the past five years, consistent with the ECDC definition of newly arrived migrants. Pooling five years of arrivals also reduces the influence of short-term migration fluctuations such as the 2022-2023 influx of Ukrainian migrants and reduced migration during the COVID-19 pandemic (2020–2021). Migration is a fluctuating phenomenon: shifts driven by conflict, climate, and other crises are intrinsic to it, thus we include migrants born in Ukraine in the main analysis while reporting their contribution separately for context (36).

Our analysis does not report on undocumented migrants, because such data are not captured in the Eurostat flows and whose omission will likely lead to underestimate the true burden.

Among HPV-attributable cancers we model cervical cancer only and do not include HPV-attributable cancers at other sites (for example anal, penile, and oropharyngeal cancers). Cervical cancer is the most important HPV-attributable cancer worldwide (80% of all HPV-attributable cancers globally are cervical (7)) and the relevant screening guidance is cervical-specific, as such excluding the other sites makes our HPV-related estimates conservative. Viral hepatitis and *H. pylori* also cause health conditions beyond cancer which are not included in the scope of this analysis, including liver cirrhosis and peptic ulcer disease (respectively).

Finally, the main analysis presents the cancers that would be averted if all migrants had access to the listed health intervention, but in reality, only a proportion of eligible individuals are offered these services, so this estimate should be considered an upper bound.

### Future research directions

Future research should address key gaps in estimating cancer burden among migrants and informing prevention policies. Real-world cancer incidence data among migrants are critical to accurately forecast health service impacts (3, 37). WHO notes that there are currently no global or regional indicators to measure the status of migrant health using routinely collected data, which can lead to a shortage of scientifically valid and comparable data on these populations (4). Implementation-oriented and community-based participatory research is important for designing effective healthcare programs. Additional analyses on efficiency and (distributional) cost-effectiveness (38) could help policymakers design effective interventions based on host country available resources and migrant population needs.

In conclusion, our analysis highlights a substantial, yet potentially avertable, burden of infection-related cancers among newly arrived migrants in the EU, with an estimated 636,000 cases expected over their lifetime. While current migrant-specific prevention guidance focuses primarily on viral hepatitis, our results support expanding to include CvC and GC prevention, alongside broader eligibility for hepatitis interventions. These services would be most effective if integrated into a comprehensive package of entry health assessments that addresses the broader health needs of newly arrived migrants, including mental health, other infectious diseases, and chronic conditions, rather than being delivered as stand-alone interventions. Our findings provide a starting point for policy discussions on cancer prevention and broader, risk-based prevention strategies tailored to migrant populations and national healthcare capacities.

## Supporting Information

### Disclaimer

Where authors are identified as personnel of the International Agency for Research on Cancer/World Health Organization, the authors alone are responsible for the views expressed in this article and they do not necessarily represent the decisions, policy or views of the International Agency for Research on Cancer /World Health Organization.

### Data availability

All data used in this analysis are publicly available. Globocan estimates are available from the International Agency for Research on Cancer’s Global Cancer Observatory (https://gco.iarc.fr//today/en). The United Nations World Population Prospects 2024 Probabilistic Projections data are available at https://www.un.org/development/desa/pd/sites/www.un.org.development.desa.pd/files/wpp2022_summary_of_results.pdf. Data on migrant population by origin, sex, and age in 2020-2024 is available from Eurostat: https://ec.europa.eu/eurostat/databrowser/product/page/migr_imm3ctb.

### Code availability

The statistical package for the ATLAS model is implemented in the methis.atlas R package, part of IARC’s METHIS modelling platform. Access to the METHIS modelling platform can be granted to a research applicant affiliated with a research institute after clearance. The clearance procedure consists of submission of a research proposal by the applicant, approval of the proposal by a dedicated access committee, and establishment of a collaborative research agreement between the research institute and IARC. More information about the model is available on the IARC’s METHIS website (https://iarc-miarc.gitlab.io/methis/methis.atlas/).

### Author contributions

CJA and IB conceived the initial project idea. CJA and RW further developed the study concept and design. RW, with assistance from DG, developed the analytical code and conducted the analyses. RW and CJA drafted the first version of the manuscript. All authors and members of the Cancer RADAR working group contributed to the refinement of the study concept and to manuscript drafting. All co-authors reviewed and approved the final version of the manuscript.

### Funding

CJA is supported by a personal VENI fellowship from the Netherlands Organisation for Health Research and Development (ZonMw; grant number 09150162210124). Additional resources, including personnel, computing infrastructure, and facilities, were provided by the International Agency for Research on Cancer of the World Health Organization (IARC-WHO). The Cancer RADAR working group contributed personal time to project idea development.

## Abbreviations

AF: Attributable fraction
ASR: Age-standardized incidence rate
CvC: Cervical cancer
ECDC: European Centre for Disease Prevention and Control
EU: European Union (27)
GC: Gastric cancer
HBV: Hepatitis B virus
HCC: Hepatocellular carcinoma
HCV: Hepatitis C virus
HIV: Human immunodeficiency virus
*H. pylori*: *Helicobacter pylori*
HPV: Human papillomavirus
IARC: International Agency for Research on Cancer
IR: Incidence rate
IQR: Interquartile range
WHO: World Health Organization
WPP: World Population Prospects
UN: United Nations

## Methods

### Overview

Below, we describe the analytical approach in detail. In brief, we combined the most up-to-date data on **(A)** the number of newly arrived migrants to countries in the EU from Eurostat (16), **(B)** cancer-specific incidence rates from the International Agency for Research on Cancer (IARC) Global Cancer Observatory (Globocan) (10), **(C)** all-cause mortality rates from the United Nations World Population Prospects (UN WPP) (56), and **(D)** the attributable fraction of each infection from previously published analyses (7, 14, 42, 49, 54) to estimate the total number of expected cancer cases over the next 80 years among migrants who immigrated to countries in the EU between 2020 – 2024. We then estimate the impact of **(E)** preventive interventions in three steps: (3a) current EU guidance on hepatitis, (3b) expanded guidance including HPV and *H. pylori* interventions, and (3c) eligibility scenarios for the hepatitis and *H. pylori* interventions. The data sources and six interventions evaluated are summarized in ***Table 1***.

### Data and assumptions

#### A. Migrant population

For this study, we used the term *newly arrived migrants* as defined by the ECDC guidance and Eurostat (6, 16). Newly arrived migrants are defined as individuals relocating to a country other than their birth country, including both intra-EU and extra-EU migrants. Eurostat provides detailed data on the age distribution, country of birth, and destination of newly arrived migrants across the 27 EU Member States (16). For this analysis, we used data on newly arrived migrants during 2020-2024. This is currently the most recent data available within the EU. Despite that, data were not always available for all EU countries. Where data were missing for age-by-birth-country, we estimated these distributions using averages derived from the aggregated migrant counts across all other EU host countries. For the host countries lacking data on the countries of birth of newly arrived migrants, we applied proportions from the 2024 UN migrant stock data. Over all five years, we had complete birth country data for 75.6% of the migrant population in this analysis. The specific countries with missing data in each year and additional details on methods to estimate missing data are available in ***Supplementary Note B.1 (Migrant population)***.

#### B. Cancer risk

The most recent (2024) age- and sex-specific cancer incidence rates (IR) for all birth countries were extracted from IARC’s Globocan database of national estimates for HCC, CvC, and GC (10). For HCC, we extracted the proportion of HCC among liver cancer from the Cancer Incidence in Five Continents (CI5) data and applied those proportions to the national liver cancer incidence estimates using a previously established approach (7, 14, 54, 57). By using birth-country (age- and sex-specific) IRs, we assume that these cancers predominantly result from infections acquired prior to migration, given the typical age at infection and the age distribution of the migrant population (58–60).

#### C. Mortality data

To account for the competing risk of mortality from all other causes in our estimation of cancer incidence over time, we applied mortality rates from the 2024 UN WPP estimates by 1-year age group, using lifetime mortality rates from each host country.

#### D. Attributable fractions

For each cancer type, we incorporated the fraction of cases attributable to each infection. For HCC, we used the birth-country-specific population attributable fraction (AF) for HBV (median 34%, IQR 13% - 54%) and HCV (median 24%, IQR 16% - 32%) (54). For GC, we used the *H. pylori* birth-country-specific population AF as calculated in a previous study using CI5 data (61). Briefly, the AF for non-cardia GC was 89% globally, and for cardia GC was typically 0% with the exceptions of east Asia (AF 29%) and China (AF 62%) (7, 42). For cervical cancer, we assumed the global fraction attributable to HPV infection was 99.7% (55).

#### E. Potential impact of prevention services upon entry

For each prevention and treatment intervention in our main analysis, we assumed all migrants who were eligible (based on sex, age, and country of birth) had full access to these services and the appropriate follow-up care was completed, acknowledging that care pathways, availability and uptake are different in each host country. Eligibility was defined based on the relevant European guidelines (e.g., recommended ages for CvC screening as per the Expert Working Group of the European Commission Initiative on Cervical Cancer) (6, 41, 42). The effectiveness of each intervention including variations by age was based on available published evidence (42, 44–53). All analysis inputs and assumptions regarding impact of prevention services for the main analysis are summarized in ***Table 1***. Lastly, it should be noted that healthcare services accessed after living in the host country for several years were not modelled in this study. For example, HPV vaccination for girls younger than 10 years old was not evaluated, as we only estimated the impact among individuals eligible at the time of arrival.

(3a) **Current EU guidance:** We modelled the three hepatitis interventions currently recommended by ECDC for newly arrived migrants: HBV vaccination, HBV screen-and-treat, and HCV screen-and-treat. ECDC recommends these for migrants from “high-risk” countries, defined as countries with an intermediate or high prevalence of chronic hepatitis B (HBsAg prevalence ≥2%) or hepatitis C (anti-HCV antibody prevalence ≥2%) (***Table 1***).

(3b) **Expanded guidance:** We added HPV vaccination, cervical screening, and *H. pylori* screen-and-treat, which are not currently part of the ECDC guidance for newly arrived migrants. In the main analysis, these were offered to all age- and sex-eligible migrants regardless of birth country (***Table 1***).

(3c) **Eligibility scenarios:** We evaluated alternative definitions of “high-risk” countries for the HCC- and GC-preventing services by identifying eligible migrants based on the age-standardized incidence rate (ASR) in their country of birth.

For the hepatitis interventions, these scenarios aim to inform discussions on widening eligibility by using the HCC ASR as an additional indicator, because as countries approach HBV and HCV elimination, current infection prevalence will increasingly underestimate the HCC burden that persists from past infections. We therefore extended eligibility as compared to the analysis in (3a), in addition to the prevalence-based criteria, to migrants born in countries in the top 25% or top 50% of HCC ASR, and to all migrants regardless of birth country. For *H. pylori* screen-and-treat, these scenarios aim to inform discussions on narrowing eligibility: compared to the analysis in (3b) wherein all migrants were eligible, in these scenarios we restricted eligibility in three scenarios: to migrants born in countries with a GC ASR ≥10 per 100,000 person-years, used to indicate populations with intermediate to high risk used by the IARC Working Group (18) (applied to sex-specific or both-sex ASRs), or in the top 50% of GC ASR. We did not evaluate country-based eligibility restrictions for HPV-targeted services, because these are already recommended for the host population and thus everyone would be eligible.

### Analysis

Using these data as inputs, we estimated the expected, infection-attributable, and potentially prevented number of HCC, CvC, and GC cases among newly arrived migrants in EU countries over the next 80 years (representing the approximate lifetime risk). To do this we used a previously published competing risk model, ATLAS, which has been described elsewhere (34, 35). In brief, the model calculates birth cohort-specific lifetime risk of cancer by using age- and sex-specific incidence rates and corresponding attributable fractions, combined with the competing risk of all-cause mortality.

We applied cancer incidence rates by sex, age, and birth country to the population of migrants aged 0-100 who arrived in countries in the EU over the most recent five years for which data are available (i.e., birth cohorts spanning 1920 – 2024). Expected cancer cases were projected over the next 80 years for each year of newly arrived migrants (2020-2024 arrivals projected to years 2100-2104 respectively). The cumulative number of cancer cases was summed across all migrants to calculate the total lifetime expected number of cases for HCC, CvC, and GC (“expected cancer cases”). We calculated the proportion of these cases that are caused by infections (“infection-attributable cases”), and then the proportion of these cases that could be avertable via the specified health services (“potentially averted cases”). We applied the effectiveness estimate of each intervention to the infection-specific IR for the applicable age groups and evaluated the potential impact on the number of cancer cases. Thus, the “potentially averted cases” that we estimated was the result of combining attributable fraction of the infectious agent, eligibility of migrants to receive the service (which varies by country of birth, age, and sex), and population effectiveness of each intervention. See ***Table 1*** for further details on the data sources, eligibility for, and effectiveness of the interventions for each cancer type. The core model calculation is summarised below. *C_total_* gives the total expected number of cancer cases and *C^prevented^* gives the number that could be averted by the modelled interventions.

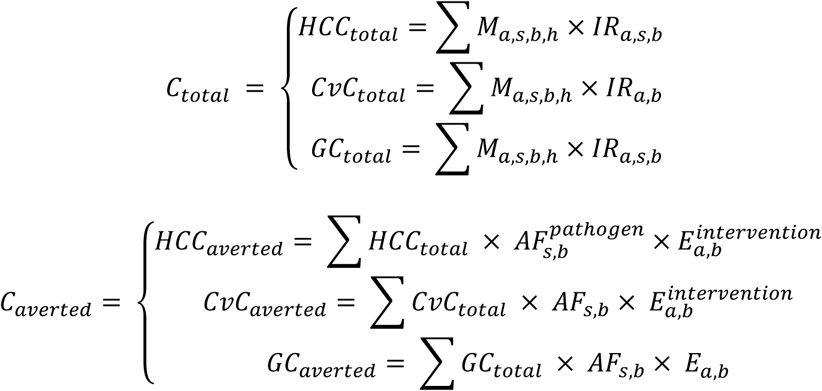

**Equation 1**. Summary of calculation approach of the main analysis. Expected (top) and averted (bottom) cancer burden among newly arrived migrants. Term definitions:, *C_total_* = total lifetime expected cancer cases among newly arrived migrants as resulting from the ATLAS competing risk model, *C_averted_* = averted cancer cases among newly arrived migrants due to the preventive services, *HCC* = hepatocellular carcinoma, *CvC* = cervical cancer, *GC* = gastric cancer, *M* = migrants ages 0-100 who arrived in countries in the EU, *IR* = cancer incidence rate, *AF^pat^*^ℎ*ogen*^ = attributable fraction of each pathogen (HCC has two causal pathogens: HBV and HCV), *E^intervention^* = effectiveness of each included preventive intervention (for cancers with multiple interventions), *a* = 2-year age group, *s* = sex, *b* = birth country, ℎ = host country.

All analyses were performed in R version 4.1.3. Access to the model code is described in the Code availability statement.

#### Sensitivity analyses

We defined several sensitivity analyses *a priori* covering (**1**) demographic parameter uncertainty (i.e., mortality rate and birth country) and, (**2**) uncertainty regarding intervention effectiveness and uptake. (**1**) We varied host country mortality rate by ±5%, a range adapted from the uncertainty intervals (UIs) provided by UN WPP (since UIs are not provided by age groups). For countries which were missing migrant arrivals data in Eurostat and stock data was used in its place (see section A. Migrant population), we defined a spread over which different possible birth country mixes could occur based on the average degree of mismatch between arrivals data and stock data for the 14 countries which have both datasets available, then drew 1000 draws of possible migrant origin mixes, calculated expected cancer cases for each mix, and reported the 2.5^th^ and 97.5^th^ percentiles of expected cancer cases as our uncertainty interval for imputed migrant birth country when missing for a specific year and host country (***Supplementary Figures S-4 and S-5***).

(**2**) The population-level effectiveness of some evaluated interventions is uncertain or context-dependent, and the degree to which migrants will realistically access these services is uncertain. We therefore assumed 50% access/uptake for each intervention (instead of 100% as done in main analysis). For CvC, we increased the population-level impact of cervical cancer screening from 60% to 90%. For H. pylori screen-and-treat, we assumed lower (40-60% effectiveness, varying by age) and optimistic (70-90% effectiveness, varying by age) effectiveness and lower effectiveness in older ages (5% in individuals older than 75). Additional details on all sensitivity analyses are provided in **Supplementary Note B.2 (Sensitivity analyses methods).**

