## Supplementary Material for "Modelling the expected and potentially avertable burden of infection-related cancer among newly arrived migrants in the European Union"

### Part A: Results Supplement

#### A.1 Additional results from main analysis

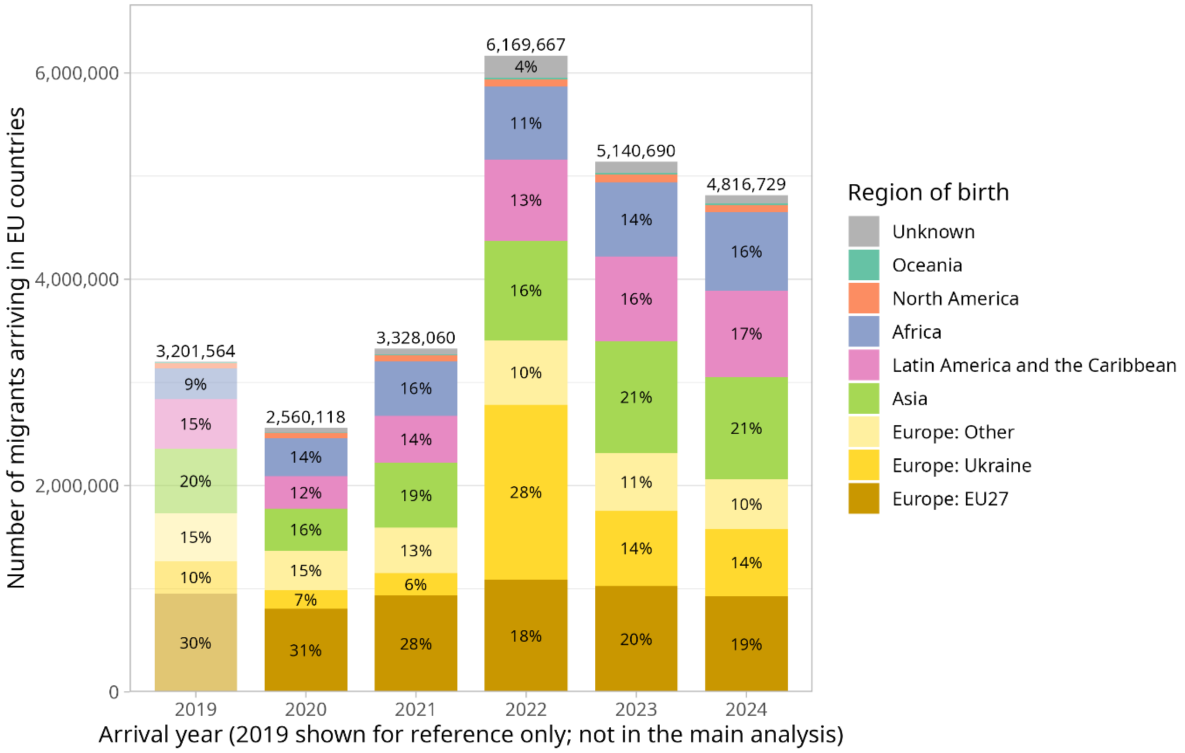

**Supplementary Figure S-1. Migrant population by year (2019-2024) and UN region of birth country.**

Each colour bar represents a UN region, with Europe subdivided into three groups: EU27 (the host countries in this analysis), Ukraine (who make up a large proportion of the migrant population), and Europe: Other (includes non-EU countries e.g., Switzerland, United Kingdom, and Russia). This analysis is conducted for migrants arriving in 2020-2024, but 2019 is shown here for comparison to understand the relative decrease in number of migrants in 2020 due to the COVID-19 pandemic. The proportion of migrants from Ukraine is also shown to illustrate the impact of the recent conflict on migration patterns during our analysis period.

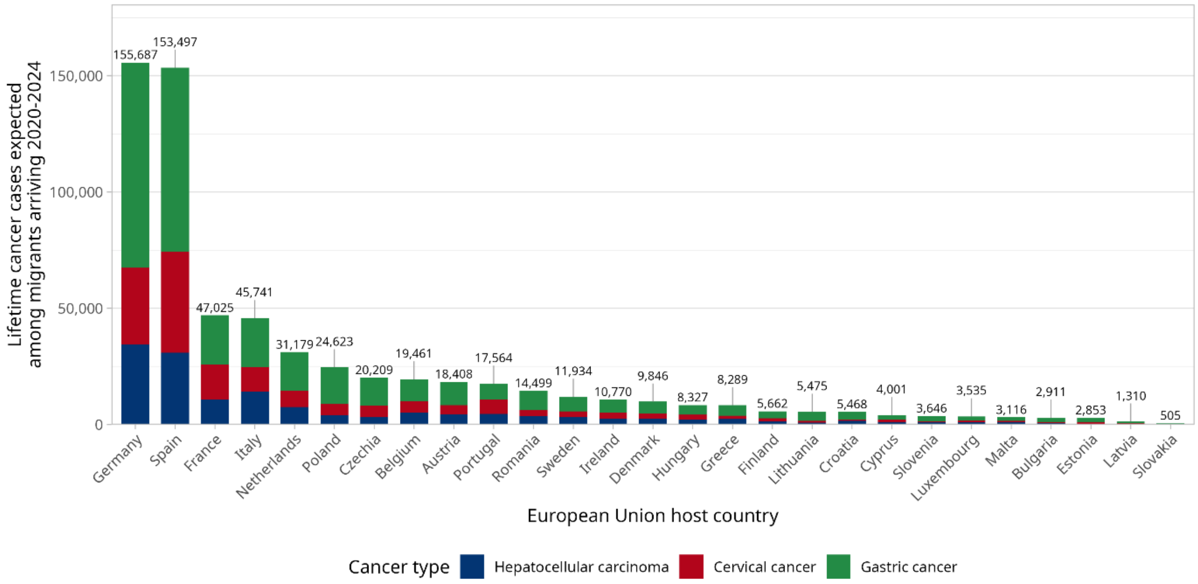

**Supplementary Figure S-2. Expected number of cancer cases among newly arrived migrants by EU host country 2020-2024.**

For each host country, the number of expected cancer cases among migrants is shown in total (label on top of each bar), and by cancer type: hepatocellular carcinoma (blue), cervical cancer (red), and gastric cancer (green). Germany and Spain have the highest number of expected cancer cases, corresponding with their rank as the countries with the largest number of migrants during the analysis period. These expected cancer cases are based on imputed migrant birth country proportions for these countries which were missing detailed arrivals data in Eurostat in at least one of the analysis years: Belgium, Cyprus, Denmark, Estonia, France, Greece, Hungary, Ireland, Malta, Poland, Portugal, Romania (see ***Supplementary Note B.1*** for more information on the missingness and imputation).

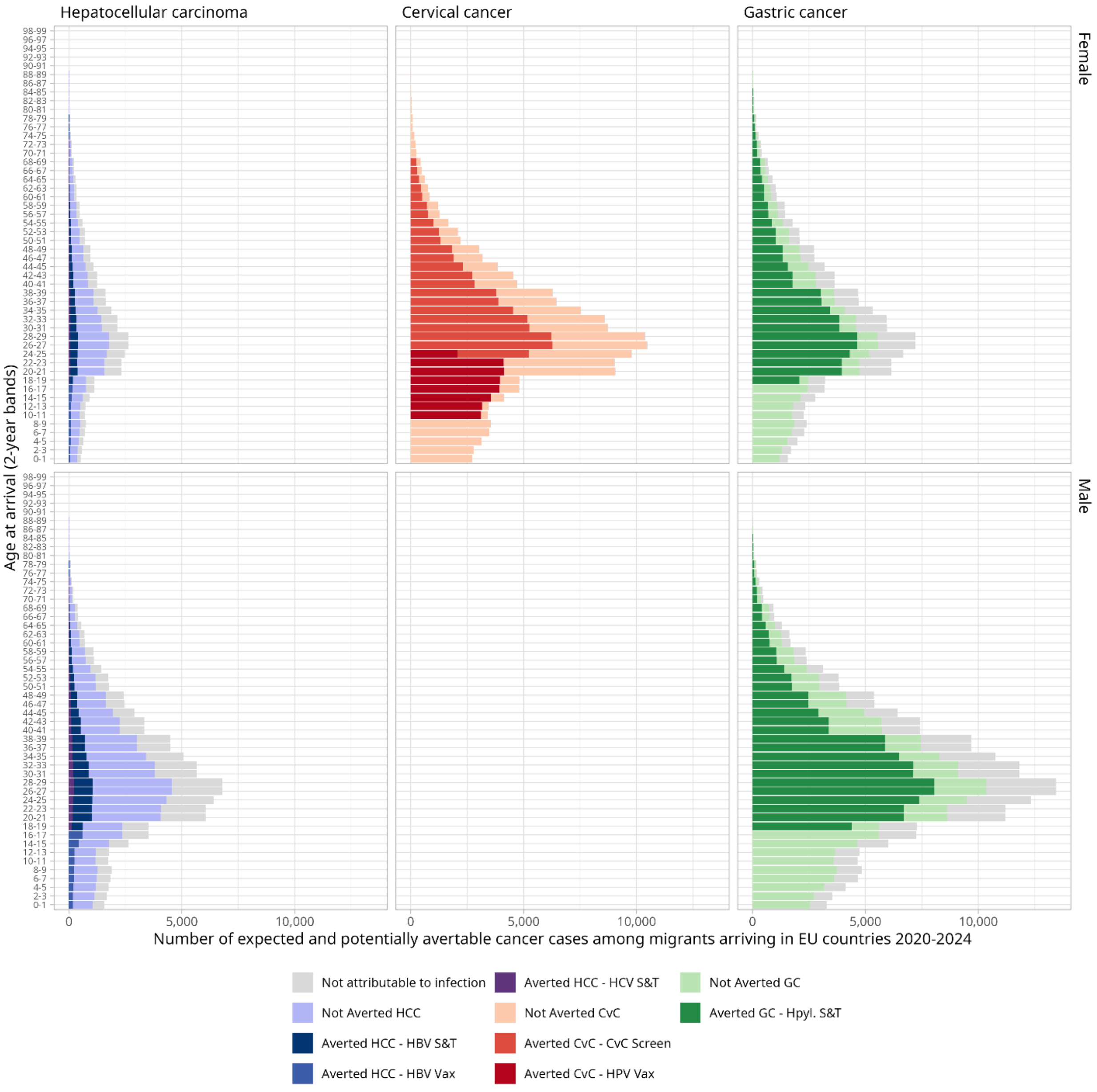

**Supplementary Figure S-3. Expected and averted cancer cases by age group, infection-related cancer and by sex among all newly arrived migrants in the EU over the most recent five years.** On the x-axis, the number of cancer cases among newly arrived migrants is shown, and on the y-axis, the age groups are displayed. The impact of the modelled interventions is illustrated for all migrants arriving 2020-2024 and therefore also includes migrants who are not eligible for the intervention upon arrival, for example, (1) girls under 10 (vaccination) and women under 25 or over 69 (screening), or (2) migrants that are not eligible for HBV or HCV testing and treatment programs based on the viral hepatitis prevalence in birth country. See ***Table 1*** for an overview of the projected interventions and eligibility criteria. Summed across all migrants, rather than just eligible migrants as presented in the main analysis, the modelled interventions would avert 14.9% (21,520/144,618) of HCC, 54.8% (84,845/154,688) of CvC, and 46.0% (154,751/336,235) of GC cases.

#### A.2 Sensitivity analyses results

The sections below discuss the results underlying ***Figure 5*** (tornado diagram) in the main manuscript in further detail.

##### A.2.1 All expected cancer cases

**Key takeaways:** Uncertainty across a plausible range of mortality rates does not have a large effect on expected cancers as compared to uncertainty for other parameters including country eligibility and service uptake. Of the three cancers, it has the largest effect on GC cases (as expected), since the age of diagnosis for that cancer is later in life than HCC and CvC, thus it is more impacted by competing mortality rates due to other causes. Uncertainty in birth country of migrants could impact the expected cancer cases by approximately -2.3% to +2.9% (***Figure 5***)

Uncertainty of birth country of migrants can affect the total number of expected cancers, though when based on a realistic range of uncertainty (as defined by comparing arrivals versus stock data for the 14 countries which have both available), the impact is minimal. **This implies that regular year-on-year fluctuations in the proportions of birth country makeup of migrants will not greatly increase or reduce the expected burden of cancer** in this population (not including such large-scale migration events such as pandemics and new inter-state conflicts).

##### A.2.2 Liver cancer (HCC)

**Key takeaways:** Applying the ECDC-recommended hepatitis prevalence threshold of 2% reduces the number of migrants eligible for viral hepatitis test-and-treat interventions. Under this criterion, migrants from 17 countries are eligible for HCV screening and treatment, and migrants from 101 countries for HBV (the 105 main analysis countries are the union of these two lists). To understand the impacts of altering these eligibility criteria, we conducted sensitivity analyses using age-standardized incidence rates (ASRs) to define eligibility for these interventions rather than prevalence.

Reconsideration of eligibility criteria is increasingly important, as the global elimination of HBV and HCV is becoming more feasible. Using ASR-based criteria for HCV (e.g., targeting migrants from the top 25%, top 50%, or all countries by ASR) resulted in a greater total reduction in HCC burden. For HBV, ASR-based criteria also produced a greater total reduction in HCC burden.

**The results of this analysis suggest that expanding the currently used 2% viral prevalence threshold to also include countries with HCC ASRs above the global median may be worth considering.** Such an approach would increase the number of cancer cases averted in the migrant population.

##### A.2.3 Cervical cancer (CvC)

**Key takeaways:** Access to screening and appropriate follow-up are key determinants of cancer prevention. Assuming that only 50% of migrants accessed the screening, 33.5% fewer total CvC cancer cases were potentially averted. If the true effectiveness of screening is closer to a 90% reduction in cervical cancer (CvC) cases, nearly three quarters (73%) of total expected CvC cases would be averted.

##### A.2.4 Gastric cancer (GC)

**Key takeaways:** Population-wide implementation of *H. pylori* screen-and-treat (i.e. offered to all migrants regardless of country of birth) is estimated to avert approximately half of the expected gastric cancer (GC) cases.

In the “optimistic effectiveness” sensitivity analysis, we assumed that future trial results will provide more precise estimates of incidence-rate reductions associated with population-based *H. pylori* screen-and-treat programmes using bismuth quadruple therapy. If optimistic effectiveness estimates are close to reality, over half (52%) of GC cases among migrants could be averted through a breath test followed by a 14-day treatment regimen for those testing positive at entry.

In sensitivity analyses evaluating more restrictive eligibility criteria for screen-and-treat, we applied an eligibility criterion broadly aligned with working group recommendations by offering *H. pylori* screen-and-treat only to migrants from countries with GC ASRs above the global median, which would avert 41% of GC cases, compared with 46% under universal eligibility. **In other words, by limiting *H. pylori* screen-and-treat to migrants from the highest-risk half of birth countries, a similar population impact could be had as under universal eligibility for migrants.**

### Part B: Methods Supplement

#### B.1 Migrant population

##### B.1.1 Migrant population calculation approach

Additional notes on the calculation of the migrant population when key data were not reported.

**Missing age distribution:**

For host countries including Austria, Estonia, Greece, Ireland, Malta, Romania, and Slovenia (full list by year specified in ***Supplementary Table S-1a,*** proportion missing data in ***Supplementary Table S-1b***) without Eurostat data on the age distribution of migrants by country of birth, we applied the average age distribution by each migrant’s country of birth observed across all other EU host countries. Slovakia was excluded from this calculation because its age distribution was markedly skewed toward younger ages (***Supplementary Table S-2***).

**Missing birth country:**

For host countries including Belgium, Cyprus, Denmark, Estonia, France, Greece, Hungary, Ireland, Malta, Poland, Portugal, and Romania (***Supplementary Table S-1a,*** proportion missing data in ***S1-b***) without Eurostat data on the country of birth of newly arrived migrants, we derived the distribution of migrants by country of birth from the 2024 UN migrant stock data and applied these proportions to the total number of newly arrived migrants in each respective host country. When host countries reported a number of migrants with “Unknown” as the country of birth, we redistributed these individuals proportionally across the observed countries of birth for migrants to that specific host country to approximate the total number of cancer cases.

Table S-1a: Missing migration data for European Union countries by year

| Year | Missing migrant age-by-birth country | Missing migrant birth country | Missing both |
|---|---|---|---|
| 2020 | Austria, Slovenia | Belgium, Cyprus, Estonia, France, Poland, Portugal | Greece, Ireland, Malta, Romania |
| 2021 | Austria, Romania, Slovenia | Belgium, Cyprus, Estonia, France, Hungary, Poland, Portugal | Greece, Ireland, Malta |
| 2022 | Austria, Estonia, Romania, Slovenia | Belgium, Cyprus, Denmark, France, Hungary, Poland, Portugal | Greece, Ireland, Malta |
| 2023 | Austria, Romania, Slovenia | Denmark, France, Hungary, Poland, Portugal | Greece, Ireland, Malta |
| 2024 | Austria, Romania, Slovenia | Denmark, France, Hungary, Poland | Greece, Ireland, Malta, Portugal |

Table S-1b: Missing migration data proportion by year as reported in Eurostat

| Year | Total foreign-born arrivals | Complete data (%) | Age imputed (%) | Birth country imputed (%) | Age and birth imputed (%) |
|---|---|---|---|---|---|
| **2020** | 2,641,523 | 66.4 | 4.8 | 21.6 | 7.2 |
| **2021** | 3,440,796 | 65.9 | 6.5 | 24.6 | 2.9 |
| **2022** | 6,319,718 | 70.6 | 6.9 | 18.9 | 3.6 |
| **2023** | 5,308,042 | 70.2 | 6.7 | 18.9 | 4.2 |
| **2024** | 4,970,939 | 70.6 | 5.6 | 16.3 | 7.5 |
| **2020-2024** | 22,681,018 | 69.3 | 6.3 | 19.5 | 4.9 |

Table S-2: Migrant population by European Union host country as used in this analysis

| Host country name | Host ISO3 | Total migrants 2020-2024* | Male (%) | Median age (IQR)** |
|---|---|---|---|---|
| Germany | DEU | 5,486,211 | 54% | 28 (19 - 39) |
| Spain | ESP | 4,518,933 | 51% | 32 (22 - 44) |
| France | FRA | 1,704,116 | 49% | 29 (22 - 39) |
| Italy | ITA | 1,487,739 | 57% | 29 (20 - 39) |
| Netherlands | NLD | 1,107,268 | 51% | 27 (20 - 36) |
| Poland | POL | 1,063,545 | 60% | 28 (16 - 41) |
| Czechia | CZE | 721,664 | 48% | 31 (19 - 42) |
| Belgium | BEL | 712,755 | 53% | 28 (20 - 38) |
| Austria | AUT | 656,815 | 52% | 29 (20 - 41) |
| Romania | ROU | 626,588 | 66% | 29 (20 - 39) |
| Portugal*** | PRT | 584,880 | 54% | 30 (22 - 40) |
| Ireland | IRL | 433,282 | 47% | 29 (19 - 40) |
| Sweden | SWE | 427,948 | 51% | 29 (20 - 38) |
| Denmark | DNK | 363,847 | 51% | 28 (21 - 38) |
| Greece | GRC | 328,695 | 59% | 29 (22 - 40) |
| Hungary | HUN | 301,351 | 58% | 30 (21 - 42) |
| Croatia | HRV | 224,418 | 72% | 34 (26 - 44) |
| Lithuania | LTU | 207,780 | 71% | 34 (25 - 43) |
| Finland | FIN | 192,863 | 54% | 27 (17 - 34) |
| Cyprus | CYP | 154,602 | 50% | 29 (20 - 40) |
| Bulgaria | BGR | 140,884 | 57% | 33 (19 - 49) |
| Malta | MLT | 134,168 | 64% | 28 (16 - 39) |
| Luxembourg | LUX | 124,562 | 53% | 29 (22 - 39) |
| Slovenia | SVN | 123,917 | 63% | 29 (21 - 40) |
| Estonia | EST | 103,339 | 51% | 30 (19 - 43) |
| Latvia | LVA | 56,846 | 53% | 32 (18 - 46) |
| Slovakia | SVK | 26,245 | 55% | 9 (3 - 34) |

Notes: *The total migrants in 2020-2024 in this table is less than the total number presented in Supplementary Table S-1b because the dataset for analysis was limited to migrants with cancer data available in Globocan, which excludes migrants from certain birth countries including European microstates (e.g., Andorra, Monaco) and small Caribbean and Pacific states (e.g., Dominica, Grenada, Micronesia). **Age is imputed for countries missing age distribution data in Eurostat: Austria, Estonia, Greece, Ireland, Malta, Romania, Slovenia .***Eurostat is missing 2024 total migrants data for Portugal, and the 2023 total was carried forward to use in its place.

#### B.1.2 Data validation approach

To measure how far the stock composition departs from the composition of recent arrivals, we used the 14 host countries that reported arrivals by country of birth in every year from 2020 to 2024 (Austria, Bulgaria, Croatia, Czechia, Finland, Germany, Italy, Latvia, Lithuania, Luxembourg, the Netherlands, Slovakia, Slovenia and Spain).

For each, we compared the observed composition of arrivals, q, with its UN stock composition, p, using the dissimilarity index, $DI = \frac{1}{2} \Sigma_i |p_i - q_i|$, summed over birth countries. The DI is the percentage of migrants who would have to be reassigned to a different birth country for the two compositions to match. It ranges from 0 (identical) to 100% (no birth countries in common).

Weighted by foreign-born arrivals, the mean DI between stock and observed arrivals was 27.7% (***Figure S-4***) . We conducted this same DI analysis to compare if pooling migrant arrivals data from all EU countries for which it was available would be a better match than using stock data for each host country. The resulting DI was higher (41.3%), indicating the stock data was a better match for the true arrivals data.

The authors are not aware of any other data sources reporting migrant arrivals by age, sex, and birth country for EU host countries, as required for this analysis. Thus, use of UN stock data is the best alternative available, and we conduct sensitivity analyses (detailed in ***Supplementary Note B.2*** below) to quantify a plausible range of uncertainty around the use of these data.

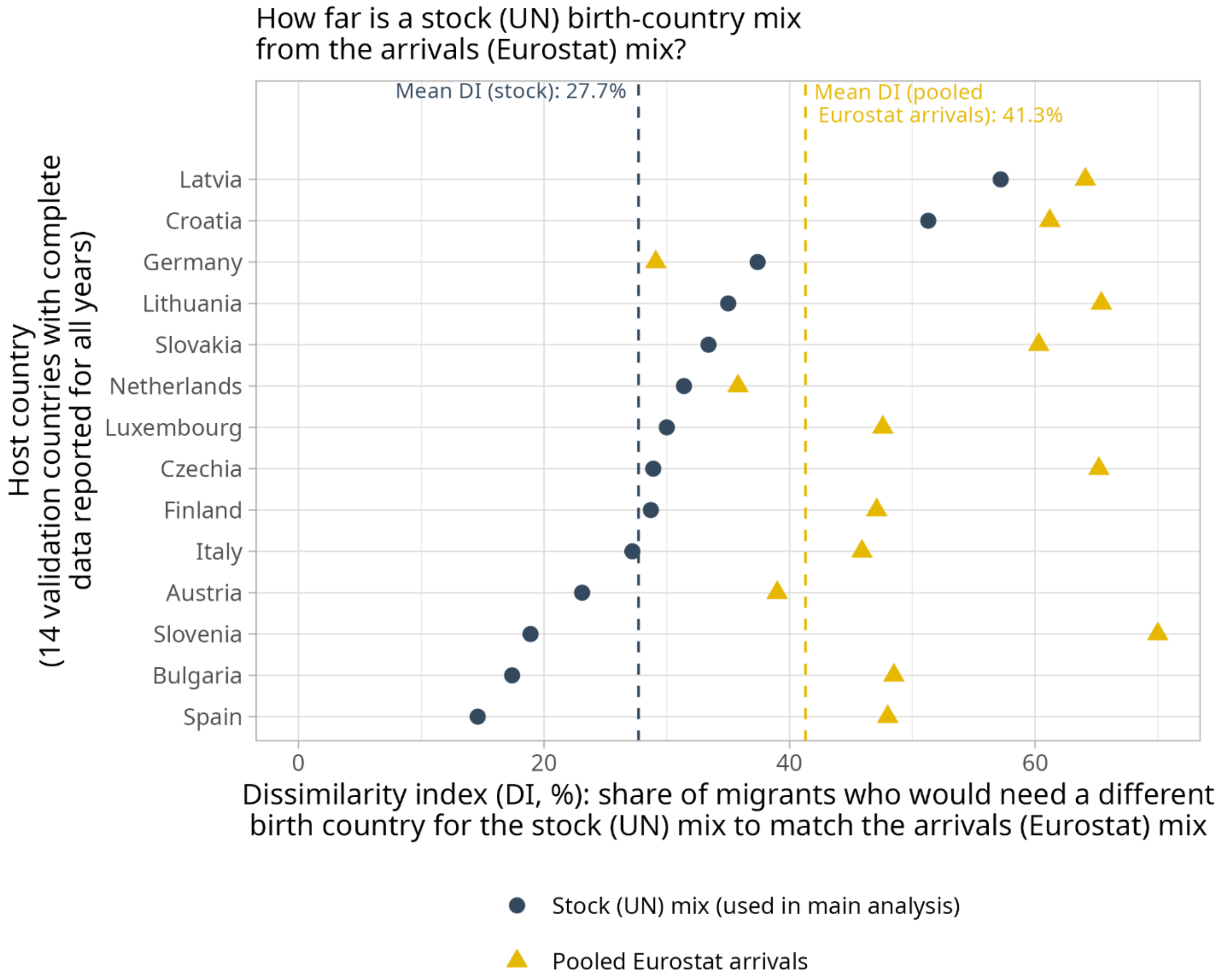

Supplementary Figure S-4. Dissimilarity index (DI) between migrant stock (UN) and migrant arrivals (Eurostat) data.

A DI of 0% would mean the data are identical, while 100% would mean no birth countries in common between the two. The DI for two approaches to imputing birth country for missing-data countries are shown: A) birth country proportions per the host country-specific migrant stock data (blue circles) B) birth country proportions per pooled Eurostat flows uniform for all European Union countries (yellow triangles). In almost all cases (except for Germany, for which the pooled EU data is the best match because Germany has the largest migrant population), the host country stock data is the closest approximation (i.e., lower DI) to the host country Eurostat arrivals data. Stock was then used for imputation and as the basis for the sensitivity analyses.

#### B.2 Sensitivity analyses methods

##### B.2.1. Demographic parameter uncertainty

We conducted two sensitivity analyses for population-wide parameters which affect the expected cancer cases: population mortality rate and migrant birth country uncertainty for the countries without migration data reported in Eurostat.

**Mortality rate uncertainty:**

UNWPP only provides 95% uncertainty intervals (UI) for mortality rate estimates for the whole population, not the age-specific values. We took the size of the UI for the total population, which was +/- 5% around the central estimate for the mortality rate for Europe as a whole, and applied to the age-specific mortality rates. Thus we analysed a deterministic high/low estimate of +/-5% around each age-specific and host-country-specific mortality rate.

**Birth country uncertainty:**

Eurostat does not report immigration by country of birth for all host countries in all years (***Table S-1a***). These host-years accounted for 24.4% of foreign-born arrivals to the EU over the study period (***Table S-1b***). In the main analysis, missing-data host countries' total foreign-born arrivals were distributed across birth countries according to each host country's 2024 migrant stock composition. Because the stock population reflects accumulated migration over several decades, it may differ from the composition of recent arrivals. We quantified this difference as a dissimilarity index (DI) (***Figure S-4***), and propagated this degree of mismatch through the model probabilistically using Dirichlet sampling.

For each host country with imputed origins, and separately by sex, we drew 1,000 birth-country compositions from a Dirichlet distribution centred on its stock composition μ, with concentration parameter $\alpha_0$ ($p \sim \text{Dirichlet}(\alpha_0\mu)$). We chose $\alpha_0$ so that the mean DI between the sampled compositions and the stock composition, weighted by migrants, equalled the 27.7% observed in the validation countries ($\alpha_0 = 45.1$). Each sampled composition was applied to all of that host country's imputed years and to all three cancer sites. A host country's male and female compositions were perturbed jointly, while host countries were sampled independently of each other. Host-years with reported birth-country data were held at their observed composition. We report the 2.5th and 97.5th percentiles of total EU migrant cases across the 1,000 samples as a 95% uncertainty interval (UI).

The 95% UIs for total expected cases among migrants arriving in 2020–2024 were −2.3% to +2.9% for cervical cancer, −1.2% to +1.3% for gastric cancer and −2.1% to +2.6% for liver cancer (HCC) (see ***Figure S-5***). An important limitation of this approach is that the validation countries are not the countries with imputed origins. If recent arrivals to the largest of these (France and Poland) differ from their long-standing stock more than in the 14 validation countries, the true uncertainty could be larger.

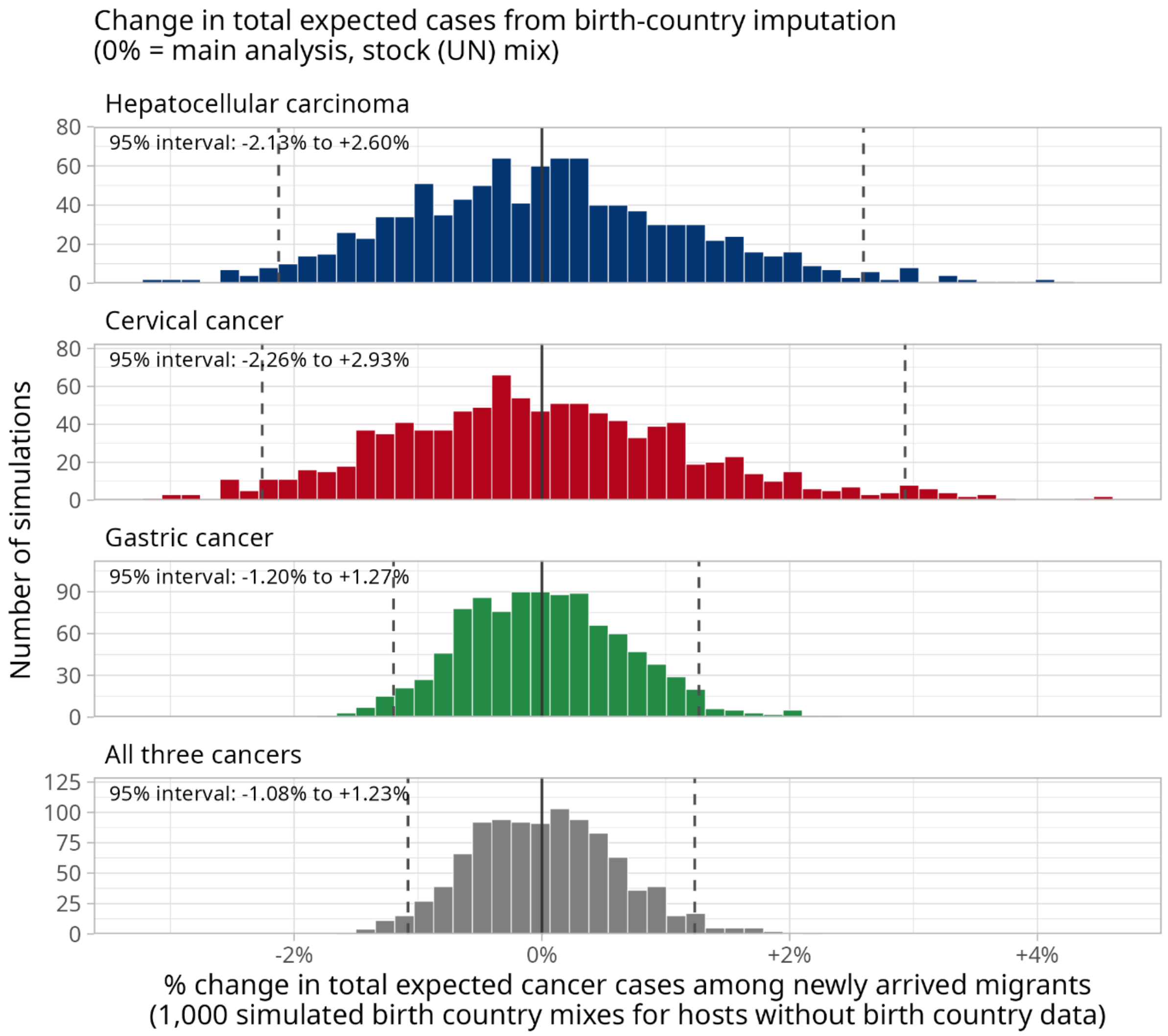

Supplementary Figure S-5. Sensitivity of total expected cancer cases to the birth-country mix assumed for host countries and years without Eurostat arrivals data by birth-country.

For each of those hosts, 1,000 alternative birth-country mixes were simulated around the migrant stock (UN) mix used in the main analysis, with the spread calibrated so that the average dissimilarity index (DI) between a simulated mix and the stock mix matches the DI observed in ***Supplementary Figure S-4*** (27.7%). Each histogram shows the resulting % change in EU-wide expected cases for HCC, CvC, GC and all three cancers combined. The solid line marks the main analysis (0%) and the dashed lines mark the 95% simulation interval, which was (-1.08%, +1.23%) for all three cancers combined.

#### B.2.2. Effectiveness and uptake uncertainty

We conducted one-way deterministic sensitivity analyses for intervention effectiveness parameters based on reviewing the literature and expert opinion. We also present results assuming only 50% of migrants had full access to the intervention (rather than 100%).

Table S-3. Sensitivity analyses of effectiveness and uptake for prevention interventions

| Cancer | Analysis | Description | New parameter values |
|---|---|---|---|
| **HCC, CvC, and GC** | **50% uptake** | Main analysis, except assuming only half of eligible migrants receive the full screen-and-treat service, whether due to lack of access, lack of confirmatory testing, lack of follow-up, or any similar issue resulting in incomplete treatment | **Access to service: 50% of eligible** |
| **CvC** | **Optimistic screening effectiveness** | Assumes 90% impact of screening programs if all migrants have high access, follow-up, and treatment. | **Ages & impact on cancer:**<br>• Age 25-69: **90%** reduction of IR |
| **GC** | **Optimistic screening effectiveness** | Main analysis, but with optimistic estimate of impact on cancer based on recent observational data from *H pylori* eradication trials (1) | **Ages & impact on cancer:**<br>• Age 18-39: **90%** reduction IR<br>• Age 40+: **70%** reduction of IR |
| | **Effectiveness as previous standard of care** | Main analysis, but with lower effectiveness estimates based on the previous standard of care. This regimen is outdated but more robust data on its effectiveness exists (2, 3) | **Ages & impact on cancer:**<br>• Age 18-39: **60%** reduction IR<br>• Age 40+: **40%** reduction of IR |
| | **Lower effectiveness in older ages** | Main analysis, but assuming lower impact in older adults aged 75+ years. The lower estimate is based on modelling data from a US-based study (4) | **Ages & impact on cancer:**<br>• Age 18-39: 80% reduction IR<br>• Age 40-74: 60% reduction of IR<br>• **Age 75+: 5%** |

#### B.2.3. Migrant eligibility scenarios

The HCC main analysis is based on ECDC guidelines which restricts eligibility for prevention services based on viral hepatitis prevalence in the migrant's birth country. We evaluated three alternative eligibility scenarios with expanded criteria for prevention services: two based on age-standardized cancer risk in the birth country and one with all migrants being eligible.

The GC main analysis is based on the IARC Gastric Cancer Working Group report which identifies migrants (in general) as a high-risk population, and we thus model as if all migrants are eligible for screen-and-treat. Population-wide guidance for screen-and-treat is evolving and is accompanied by concerns of inefficient use of resources in resource-constrained settings and possible consequences for antimicrobial resistance. Accordingly, we present three more restrictive eligibility scenarios based on age-standardized incidence rates of GC in the birth country.

No eligibility scenarios were evaluated for CvC prevention. Unlike for GC and HCC, the proposed vaccination and screening interventions for migrants in this analysis are the same guidelines which are recommended for host country populations. Thus, variations of eligibility criteria are not being considered.

Table S-4. Sensitivity analyses of migrant eligibility scenarios for HCC and GC prevention

| Cancer | Analysis | Description | Impact on eligible countries |
|---|---|---|---|
| **HCC** | high-risk countries definition 1: top 25% HCC ASR | Migrants from countries in the top 25% of ASR (either sex) for HCC | Birth countries eligible: 119 (14 more than main analysis) |
| | high-risk countries definition 2: top 50% HCC ASR | Migrants from countries in the top 50% of ASR (either sex) for HCC | Birth countries eligible: 146 (41 more than main analysis) |
| | high-risk countries definition 3: All countries eligible | Migrants from any birth country are eligible for prevention services | Birth countries eligible: All (185 countries in Globocan) |
| **GC** | high-risk countries definition 1: both-sex GC ASR >= 10 | Main analysis, restricted to high-risk countries defined as those with overall ASR ≥ 10 per 100,000 person-years (3) | Birth countries eligible: 42 (143 fewer than main analysis) |
| | high-risk countries definition 2: sex-specific GC ASR >= 10 | Main analysis, restricted to high-risk countries defined as those with either sex-specific ASR ≥ 10 per 100,000 person-years | Birth countries eligible: 61 (124 fewer than main analysis) |
| | high-risk countries definition 3: top 50% GC ASR | Main analysis, restricted to high-risk countries defined as those with ASR in the top 50% (either sex) of all countries globally | Birth countries eligible: 105 (80 fewer than main analysis) |
